# Repeated CHARGE-AF Score Assessments for Predicting Incident Atrial Fibrillation: A Longitudinal Analysis from the ARIC Cohort

**DOI:** 10.64898/2026.07.10.26357684

**Authors:** Eduardo Gonzalez Villarreal, Faye Norby, Lin Yee Chen, Linzi Li, Oluseye Ogunmoroti, Louis Y. Li, Elsayed Soliman, Alvaro Alonso

## Abstract

**Background:** The Cohorts for Heart and Aging Research in Genomic Epidemiology – Atrial Fibrillation (CHARGE-AF) score is a validated tool for estimating 5-year risk of atrial fibrillation (AF). We aimed to evaluate the utility of repeated CHARGE-AF scores for improving AF risk prediction.

**Methods:** We analyzed participants from the Atherosclerosis Risk in Communities (ARIC) study with complete data from the first four clinic visits (9-year period) and with no prevalent AF by visit 4 (analysis baseline; N = 10,188). CHARGE-AF scores were calculated for each visit using clinical and demographic variables. Incident AF was determined from electrocardiograms, hospital discharge codes, and death certificates over a median follow-up of 19.5 years. Four Cox regression models were assessed: model 1 included only the visit 4 CHARGE-AF score, and subsequent models added prior CHARGE-AF scores in stepwise fashion. C-statistics were used to evaluate model discrimination, and comparison of observed versus predicted risk was employed to evaluate calibration. Secondary analysis restricted follow-up to five years.

**Results:** During follow-up, 2,519 participants developed AF (14.2 cases per 1,000 person-years). The mean age of participants at start of follow-up was 62.8 (±5.6) years. In the primary analysis, each 1% increase in the visit 4 CHARGE-AF score was associated with incident AF (Model 1 HR = 1.14, 95% CI 1.13-1.15). Addition of scores from prior visits did not significantly improve model discrimination (C-statistic: 0.702-0.703 for all models). Sex modified the association between a 1% increase in CHARGE-AF score and incident AF, with a stronger association among females (Model 1 HR = 1.21, 95% CI: 1.19-1.22) than among males (HR for model 1 = 1.12, 95% CI: 1.11-1.13). Similar patterns were observed in the secondary (5-year restricted) analysis.

**Conclusions:** A single measurement of the CHARGE-AF score provided strong predictive value for incident AF, with the addition of prior scores offering limited incremental benefit. These findings suggest that, in clinical settings with longitudinal data, the most recent assessment is sufficient for AF risk prediction.

## Introduction

Atrial fibrillation (AF) is the most commonly recorded cardiac arrhythmia. It not only affects millions of adults directly, but it also contributes to stroke, heart failure, and elevates a person’s overall cardiovascular risk profile. (1,2). The prevalence of AF increases with age as 5.9% of adults over the age of 65 live with AF, and nearly 70% of all AF cases occur from ages 65 to 85. (3) AF also contributes substantially to healthcare utilization and the need for long-term management. To reduce the burden of AF both at the individual and population levels, early detection of high-risk individuals becomes essential. Risk prediction models are a critical tool in the medical context because they integrate clinical and demographic variables to identify patients at higher risk before AF even develops.

The Cohorts for Heart and Aging Research in Genomic Epidemiology – Atrial Fibrillation (CHARGE-AF) score is a validated multivariable prediction tool designed to quantify an individual’s five-year risk of incident AF (given as a percentage). (4) This score incorporates age, race, weight, height, systolic and diastolic blood pressure, use of antihypertensive medication, smoking, diabetes, prevalent coronary heart disease, and prevalent heart failure. (4) Prior studies have relied on a single timepoint of data to calculate one CHARGE-AF score per patient. (5–7) However, longitudinal risk assessment would capture any changes in clinical status that modify AF risk over time. Thus, building models that incorporate longitudinal risk scores may improve prediction for incident AF. The Atherosclerosis Risk in Communities (ARIC) study provides a unique opportunity to investigate repeated CHARGE-AF scores, given the comprehensive follow-up data available from clinical visits that now span over three decades. (8)

In this study, we evaluated the predictive utility of repeated CHARGE-AF scores (taken from four distinct ARIC visits) for incident AF. We hypothesized that the most recent CHARGE-AF score would have the strongest association with incident AF, given that components of this score reflect the latest available cardiovascular risk profile. We also hypothesized that including prior risk scores would provide very limited additional predictive value. By comparing various models, we aimed to understand the importance of repeated risk assessment for AF prediction and to inform current practices on the frequency of risk evaluation.

## Methods

### Study population

The ARIC study is prospectively following a cohort of 15,792 adults, with follow-up beginning in 1987. Participants of ages 45 to 64 were recruited from four U.S communities: Forsyth County, North Carolina; Jackson, Mississippi; suburban Minneapolis, Minnesota; and Washington County, Maryland. They have been followed longitudinally to assess cardiovascular disease risk. (9) This analysis used data from participants with complete measurements from visits 1 through 4 of the ARIC study (Visit 1: 1987–89; Visit 2: 1990–92; Visit 3: 1993–95; Visit 4: 1996–98). (8) Data collection includes measurements obtained during clinic visits and through telephone interviews. Participants were excluded if they had prevalent AF detected at or before visit 4. Outcome data is continuing to be collected. The study protocol was approved by institutional review boards at each field center. All participants provided written informed consent.

### CHARGE-AF calculation

For each eligible participant, CHARGE-AF scores were calculated at each visit using ARIC variables for age, race, weight, height, systolic and diastolic blood pressure, use of antihypertensive medication, smoking, diabetes, prevalent coronary heart disease, and prevalent heart failure. (4) Height, weight, blood pressure, and fasting glucose were measured during in-person clinic visits. Blood pressure was calculated as the average of the last two measurements obtained with a sphygmomanometer while the participant was seated. Antihypertensive medication use was determined through self-report and the verification of prescription bottles. Diabetes was defined by a fasting glucose of ≥ 126 mg/dL (or non-fasting glucose of ≥ 200 mg/dL), a self-reported physician diagnosis of diabetes, or current use of diabetes medication. Prevalent heart disease and heart failure were identified using a combination of self-report from visit 1, medical history, and hospital discharge codes (ICD-9-CM: 428.x). (10) Race and current smoking status were self-reported. (11) Race was categorized as White and non-White (non-White included participants self-reporting Black or Other race) for purposes of the CHARGE-AF calculation.

### AF ascertainment

AF was identified directly from electrocardiograms (ECGs), hospital discharge codes (ICD-9-CM: 427.31 or 427.32, ICD-10-CM: I48.x) excluding cases occurring in the context of cardiac surgery, and from death certificates listing AF as an underlying or contributing cause of death (ICD-10: I48). (12) A 12-lead ECG was recorded and transmitted to the ARIC Central ECG Reading Center for interpretation; a trained cardiologist visually inspected each ECG to confirm the diagnosis. The diagnosis of AF was primarily determined by ECG and supplemented by hospital discharge codes and death certificates. The earliest detected AF event among the three methods was used to establish the date of incident AF. Incident AF was ascertained during follow-up after Visit 4, which served as baseline for this analysis. Using all three methods in combination, AF ascertainment achieved high sensitivity and specificity in White individuals (85% and 99%, respectively) and also in Black individuals (80% and 99%, respectively). (13)

### Statistical analysis

Cox proportional hazards regression was used to evaluate the utility of CHARGE-AF scores in predicting incident AF using SAS Version 9.4; SAS Institute, Cary, NC). Follow-up time was calculated as the time elapsed from visit 4 date to AF incidence, death, loss to follow-up, or December 31, 2022 (whichever occurred first). Model 1 included CHARGE-AF score at visit 4 as the sole independent variable.

Subsequent models added previous CHARGE-AF scores (measured at visits 3, 2, and 1) in a stepwise fashion; each model contained one additional CHARGE-AF score when compared to the previous model. Model discrimination was assessed using the C-statistic, an established method to measure a model’s ability to distinguish between individuals with and without the event of interest. (14) To provide a comprehensive assessment of predictive performance, a calibration plot was generated to examine the predictive ability of CHARGE-AF over a five-year timeframe. (15) Additional analysis tested for interaction by sex and stratified Cox proportional hazards models to examine potential differences in predictive ability of CHARGE-AF for males versus females.

As a secondary analysis, the models were repeated while restricting follow-up time to five years after visit 4 date. This was done to align with the original design of the CHARGE-AF score, which was to specifically predict an individual’s five-year risk of AF. (16)

## Results

From the original ARIC cohort (N = 15,792), analysis was restricted to those participants who attended all of the first four ARIC visits, as this information was necessary to calculate CHARGE-AF scores. Because incident AF was the outcome of interest, participants with AF detected at or before visit 4 were also excluded. The progression to the final analytic sample (N = 10,188) is shown in **Figure 1**. **Table 1** presents the sample characteristics at baseline. The majority of the study sample was White (79%), with 14% of participants reporting that they were current smokers. Because our study baseline was the fourth ARIC visit, participants were on average already in their early-to-mid 60s (62.8 ± 5.6 years) at the start of the follow-up period. Participants were followed for a median time of 19.5 years (Q1-Q3: 12.2-23.7 years) from ARIC visit 4. During that follow-up window, 2,519 incident AF cases were recorded, corresponding to an incidence rate of 14.2 cases per 1,000 person-years. Median CHARGE-AF risk at visit 4 was 2.24% (Q1-Q3: 1.24-4.08%); the scores were right-skewed, indicating that most individuals were at low risk for developing AF.

**Figure 1.**
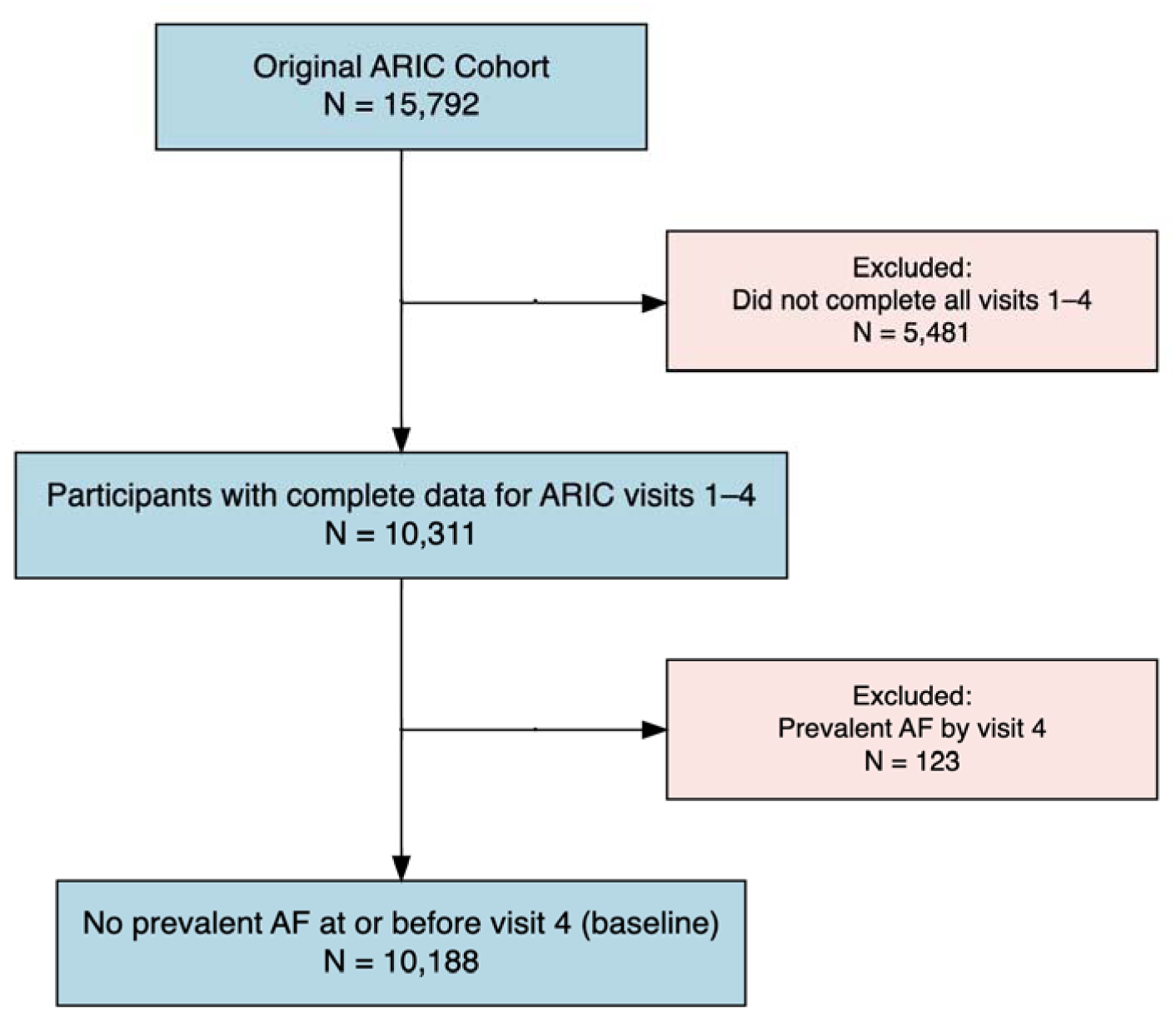
Flow of Participants from Original ARIC Cohort to Final Analytic Sample

**Table 1.** Baseline Participant Characteristics, ARIC Visit 4. (**1996–1998**)

| **Characteristic** | **Overall**<br>N = 10,188<sup>1</sup> | **Females**<br>N = 5,738<sup>1</sup> | **Males**<br>N = 4,450<sup>1</sup> |
| --- | --- | --- | --- |
| **Race** |  |  |  |
| White | 8,085 (79%) | 4,383 (76%) | 3,702 (83%) |
| Black | 2,074 (20%) | 1,340 (23%) | 734 (16%) |
| Other<sup>2</sup> | 29 (0.3%) | 15 (0.3%) | 14 (0.3%) |
| **Age (years)** | 62.8 (5.6) | 62.5 (5.6) | 63.1 (5.7) |
| **Height (cm)** | 168.4 (9.2) | 162.3 (5.9) | 176.1 (6.5) |
| **Weight (kg)** | 80.6 (17.2) | 75.4 (17.0) | 87.3 (14.8) |
| **Systolic blood pressure (mmHg)** | 127.1 (18.8) | 127.5 (19.4) | 126.6 (18.1) |
| **Diastolic blood pressure (mmHg)** | 70.8 (10.2) | 69.9 (10.2) | 72.0 (10.1) |
| **Current smoker** | 1,471 (14%) | 794 (14%) | 677 (15%) |
| **On antihypertensives** | 4,282 (42%) | 2,431 (42%) | 1,851 (42%) |
| **Diabetes** | 1,604 (16%) | 836 (15%) | 768 (17%) |
| **Prevalent heart failure** | 480 (4.7%) | 307 (5.4%) | 173 (3.9%) |
| **Prevalent coronary heart disease** | 676 (6.6%) | 226 (3.9%) | 450 (10%) |
| **CHARGE-AF 5-year risk (%)** | 2.2 (1.2-4.1) | 1.8 (1.0-3.2) | 3.0 (1.7-5.4) |

<sup>1</sup>Continuous variables are expressed as mean (SD), except CHARGE-AF 5-year risk (%) which is reported as median (Q1, Q3).
<sup>2</sup>“Other” includes participants self-identifying as Asian, American Indian or Alaskan Native, Hawaiian or Pacific Islander, or mixed-race.

Results for model 1 show that a 1% increase in CHARGE-AF risk score was associated with a 14% increased risk of AF (HR = 1.14, 95% CI: 1.13-1.15). When all four CHARGE-AF scores were included in a model (model 4), only CHARGE-AF at visit 4 (HR = 1.07, 95% CI: 1.05-1.09) and at visit 3 (HR = 1.06, 95% CI: 1.02-1.10) were associated with an increased risk of AF. The previous risk scores (from visits 1 and 2) were no longer meaningfully associated with AF after adjustment for the more recent scores. Because scores were calculated from the same individuals, CHARGE-AF at visit 4 was strongly correlated with all scores from earlier visits (visit 3: r = 0.92, visit 2: r = 0.89, and visit 1: r = 0.87). **Table 2** shows the hazard ratios for all Cox regression models.

**Table 2.** Hazard Ratios associating CHARGE-AF Score and Incident Atrial Fibrillation.

| Predictor | Model 1<sup>1</sup> | Model 2<sup>1</sup> | Model 3<sup>1</sup> | Model 4<sup>1</sup> |
| --- | --- | --- | --- | --- |
| **Primary Analysis<sup>2</sup>** |  |  |  |  |
| CHARGE-AF score at visit 4 | 1.14 (1.13-1.15),<br>p<0.001 | 1.08 (1.06-1.10),<br>p<0.001 | 1.07 (1.05-1.09),<br>p<0.001 | 1.07 (1.05-1.09),<br>p<0.001 |
| CHARGE-AF score at visit 3 | - | 1.10 (1.07-1.14),<br>p<0.001 | 1.06 (1.02-1.10),<br>p=0.002 | 1.06 (1.02-1.10),<br>p=0.004 |
| CHARGE-AF score at visit 2 | - | - | 1.09 (1.03-1.14),<br>p=0.001 | 1.07 (1.00-1.14),<br>p=0.054 |
| CHARGE-AF score at visit 1 | - | - | - | 1.03 (0.95-1.12),<br>p=0.477 |
| **Secondary Analysis<sup>2</sup>** |  |  |  |  |
| CHARGE-AF score at visit 4 | 1.11 (1.10-1.13),<br>p<0.001 | 1.07 (1.03-1.11),<br>p<0.001 | 1.05 (1.01-1.09),<br>p=0.017 | 1.05 (1.01-1.09),<br>p=0.017 |
| CHARGE-AF score at visit 3 | - | 1.07 (1.00-1.14),<br>p=0.038 | 0.96 (0.88-1.05),<br>p=0.418 | 0.96 (0.88-1.05),<br>p=0.409 |
| CHARGE-AF score at visit 2 | - | - | 1.23 (1.10-1.36),<br>p<0.001 | 1.22 (1.08-1.38),<br>p=0.002 |
| CHARGE-AF score at visit 1 | - | - | - | 1.01 (0.86-1.19),<br>p=0.856 |

<sup>1</sup> Model 1 includes CHARGE-AF score measured at visit 4; Model 2: CHARGE-AF scores measured at visits 4 and 3; Model 3: CHARGE-AF scores measured at visits 4, 3 and 2; Model 4: CHARGE-AF scores measured at visit 4, 3, 2 and 1.
<sup>2</sup>Primary analysis: full follow-up from visit 4 through end of study. Secondary analysis: follow-up limited to five years after visit 4.

A secondary analysis was conducted using the same models but now limiting follow-up time to a maximum of five years after visit 4 date – matching the prediction window of the original CHARGE-AF model (see **Table 2**). During these five years, there were 298 incident cases of AF, corresponding to an incidence rate of 6.1 cases per 1,000 person-years. Model 1 showed that a 1% increase in baseline CHARGE-AF risk was associated with an 11% increased risk of developing AF in the next five years (HR = 1.11, 95% CI: 1.10-1.13). When previous CHARGE-AF scores were added to the model, the relationship between baseline CHARGE-AF and AF was attenuated. For example, model 3 (which included the CHARGE-AF score from visits 2 through 4) showed that a 1% increase in baseline CHARGE-AF created only a 5% increased risk of AF within the next five years (HR = 1.05, 95% CI: 1.01-1.10).

**Table 3** shows the C-statistic for each model as well as the change in the C-statistic from one model to the next. Model 1, including visit 4 CHARGE-AF score only, showed good discriminative ability for AF (C = 0.702, 95% CI: 0.692-0.712). Addition of CHARGE-AF scores measured at prior visits resulted in minimal improvements in discrimination. **Figure 2** shows the calibration of the CHARGE-AF score from visit 4, with the predicted five-year AF risk on the x-axis and observed risk on the y-axis. The predicted risk in each decile matches up closely with the observed risk, suggesting that CHARGE-AF is a well-calibrated predictor of AF in the short-term. In the higher-risk deciles, there was a slight tendency for predicted risk to overestimate the observed five-year incidence of AF.

**Figure 2.**
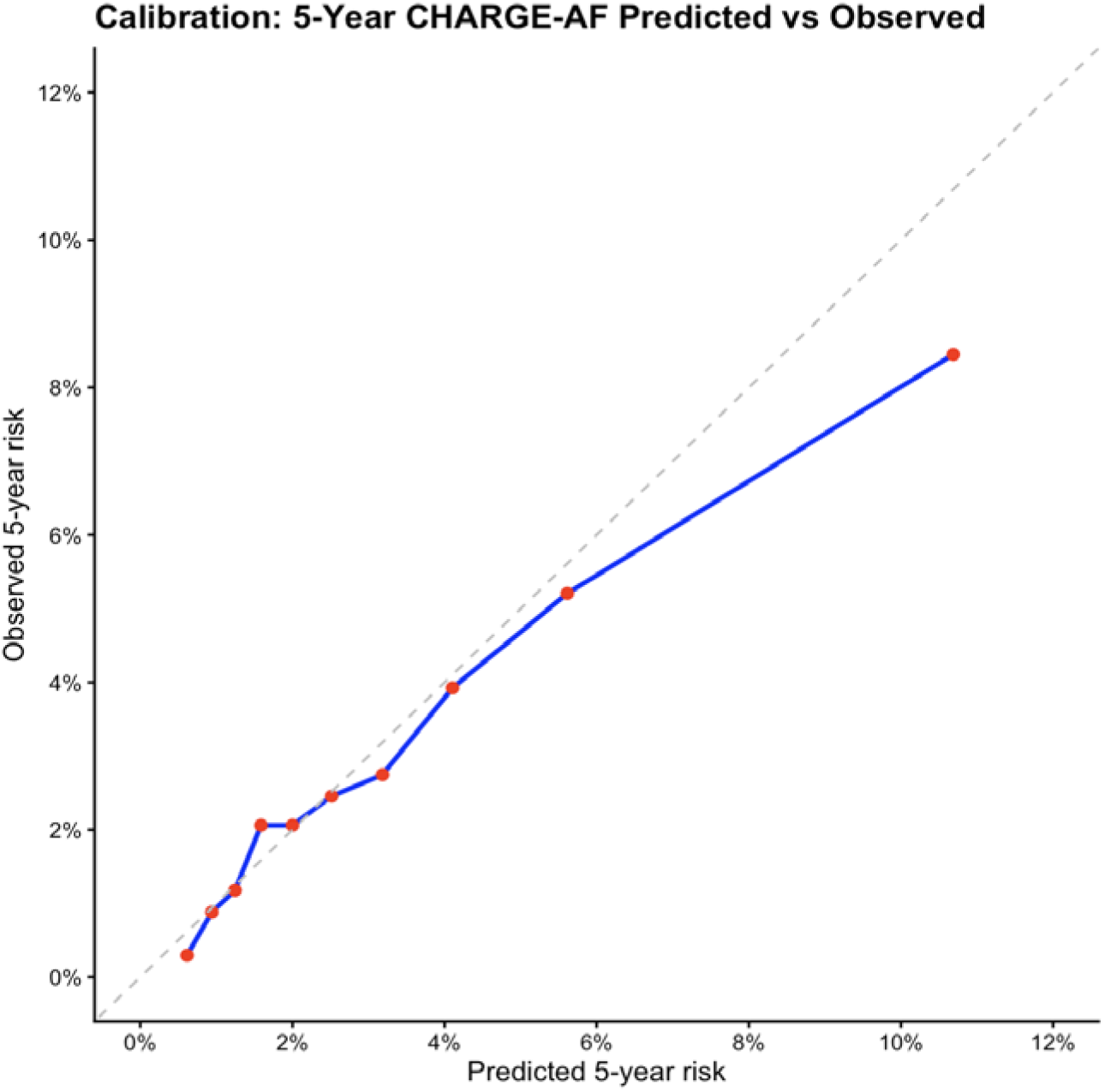
**Calibration Plot for Predicted 5-Year Atrial Fibrillation Risk**

**Table 3.** C-statistics for Cox Proportional Hazards Models*^1^*

| Model | C-Statistic | Lower 95% CI | Upper 95% CI | Increment |
| --- | --- | --- | --- | --- |
| Model 1 | 0.702 | 0.692 | 0.712 | NA |
| Model 2 | 0.703 | 0.693 | 0.713 | 0.001 |
| Model 3 | 0.703 | 0.693 | 0.713 | <0.001 |
| Model 4 | 0.703 | 0.693 | 0.713 | <0.001 |

<sup>1</sup>Models 1 through 4 are Cox regression models using CHARGE-AF scores as predictors of incident atrial fibrillation. Model 1 includes CHARGE-AF score measured at visit 4; Model 2: CHARGE-AF scores measured at visits 4 and 3; Model 3: CHARGE-AF scores measured at visits 4, 3 and 2; Model 4: CHARGE-AF scores measured at visit 4, 3, 2 and 1.

We observed a statistically significant interaction between CHARGE-AF risk score and sex (p < 0.001) for all primary models, with the effect between risk score and incident AF attenuated in males when compared to females (**Table 4**). When baseline CHARGE-AF score is the sole predictor in the model (model 1), a 1% increase in that score was associated with a 21% increased risk for AF among females (HR = 1.21, 95% CI: 1.19-1.22). In contrast, that same 1% increase in CHARGE-AF score was associated with only a 12% increased AF risk among males (HR = 1.12, 95% CI: 1.11-1.13). A similar pattern was observed across all four models, with an increase in baseline CHARGE-AF score being more strongly associated with incident AF in females. Even after stratifying by sex, model AF discriminative ability did not change when additional CHARGE-AF risk scores were included (**Table 5**).

**Table 4.** Hazard Ratios associating CHARGE-AF Score and Incident Atrial Fibrillation (Stratified by Sex)

| Predictor | Model 1<sup>1</sup> | Model 2<sup>1</sup> | Model 3<sup>1</sup> | Model 4<sup>1</sup> |
| --- | --- | --- | --- | --- |
| Male |  |  |  |  |
| CHARGE-AF score at visit 4 | 1.12 (1.11-1.13), p=<0.001 | 1.07 (1.04-1.09), p=<0.001 | 1.07 (1.04-1.09), p=<0.001 | 1.06 (1.03-1.08), p=<0.001 |
| CHARGE-AF score at visit 3 | - | 1.09 (1.05-1.13), p=<0.001 | 1.05 (1.00-1.10), p=0.035 | 1.05 (1.00-1.10), p=0.046 |
| CHARGE-AF score at visit 2 | - | - | 1.08 (1.02-1.15), p=0.009 | 1.07 (0.99-1.16), p=0.091 |
| CHARGE-AF score at visit 1 | - | - | - | 1.02 (0.92-1.13), p=0.689 |
| Female |  |  |  |  |
| CHARGE-AF score at visit 4 | 1.21 (1.19-1.22), p=<0.001 | 1.15 (1.11-1.20), p=<0.001 | 1.14 (1.09-1.19), p=<0.001 | 1.14 (1.10-1.19), p=<0.001 |
| CHARGE-AF score at visit 3 | - | 1.07 (1.02-1.13), p=0.009 | 1.04 (0.97-1.11), p=0.290 | 1.04 (0.97-1.12), p=0.271 |
| CHARGE-AF score at visit 2 | - | - | 1.09 (0.96-1.24), p=0.165 | 1.08 (0.97-1.20), p=0.137 |
| CHARGE-AF score at visit 1 | - | - | - | 0.98 (0.83-1.15), p=0.774 |

<sup>1</sup> Model 1 includes CHARGE-AF score measured at visit 4; Model 2: CHARGE-AF scores measured at visits 4 and 3; Model 3: CHARGE-AF scores measured at visits 4, 3 and 2; Model 4: CHARGE-AF scores measured at visit 4, 3, 2 and 1.

**Table 5.** **C-statistics for Sex-stratified Cox Proportional Hazards Models*^1^*** Corresponding male and female models shown side by side

| Model | Male |  |  | Female |  |  |
| --- | --- | --- | --- | --- | --- | --- |
|  | C-Statistic | Lower 95% CI | Upper 95% CI | C-Statistic | Lower 95% CI | Upper 95% CI |
| Model 1 | 0.695 | 0.680 | 0.710 | 0.697 | 0.683 | 0.711 |
| Model 2 | 0.696 | 0.681 | 0.711 | 0.698 | 0.684 | 0.712 |
| Model 3 | 0.697 | 0.682 | 0.712 | 0.699 | 0.685 | 0.713 |
| Model 4 | 0.696 | 0.681 | 0.711 | 0.698 | 0.684 | 0.713 |

<sup>1</sup>Models 1–4 are Cox regression models using CHARGE-AF scores as predictors of incident atrial fibrillation, with each subsequent model including an additional prior visit score. Model 1 includes CHARGE-AF score measured at visit 4; Model 2: CHARGE-AF scores measured at visits 4 and 3; Model 3: CHARGE-AF scores measured at visits 4, 3 and 2; Model 4: CHARGE-AF scores measured at visit 4, 3, 2 and 1.

## Discussion

This analysis of the large community-based ARIC cohort found that a single measurement of the CHARGE-AF score is adequate for the long-term prediction of AF, with addition of prior CHARGE-AF score measurements adding limited value to prediction. The Cox proportional hazards models showed that the baseline CHARGE-AF score (taken from ARIC visit 4) is a strong predictor of incident AF. This finding supports the established literature on the utility of CHARGE-AF as a tool for AF risk assessment. (4) For models with repeated measures of CHARGE-AF (models 2-4), the most recent score contributed the greatest predictive information. As CHARGE-AF scores from prior visits were added into the model, the hazard ratio for baseline CHARGE-AF was attenuated. This attenuation suggests that prior CHARGE- AF scores provide overlapping information rather than an independent predictive contribution, as reflected by their strong correlations.

The secondary analysis (limiting follow-up time to five years after visit 4) showed a weaker association between CHARGE-AF risk and incident AF, though this association was still statistically significant. Restricting follow-up reduces statistical precision; fewer AF events occurred. Similar to results from the primary analysis, the addition of past risk scores reduced the strength of the association between baseline CHARGE-AF and AF. Notably, the models’ ability to predict AF was not meaningfully different over the short-term (five years) versus the full follow-up window, suggesting that CHARGE-AF retains utility for longer-term risk prediction.

The use of the CHARGE-AF score was originally validated in three prospective cohorts (including the ARIC cohort) to ensure its generalizability to populations with diverse cardiovascular risk profiles (4). Prior studies have also found that CHARGE-AF outperforms other common clinical risk scores (such as CHA_₂_DS_₂_-VASc) for predicting incident AF. (17) Our analysis builds on past research by examining repeated risk measurements on the same participants over time – thus adding a longitudinal component to risk prediction. Our findings show that the most recent CHARGE-AF score was most strongly associated with incident AF, and that previous risk scores do not meaningfully enhance risk prediction. Clinically, these findings highlight the importance of wellness visits; they allow healthcare providers to create patients’ AF risk profile and identity high-risk individuals before cardiovascular complications arise.

Evaluation of the C-statistics shows that the discrimination is very similar across all models, even after stratifying by sex, with substantial overlap of confidence intervals across all models and minimal increments in the C-statistic. This suggests that addition of prior CHARGE-AF scores does not improve prediction beyond the information provided by the most recent assessment. Therefore, a single CHARGE-AF score is sufficient to inform an individual’s risk of developing AF. While past risk scores may provide additional clinical insight into a patient’s health trajectory, the most recent score (defined as ARIC visit 4 in our study) appears adequate as a stand-alone metric to predict AF risk. Calibration analyses further demonstrate the utility of the CHARGE-AF score in predicting AF in a five-year timeframe. Because discrimination and calibration assess distinct aspects of performance, the Cox regression model may correctly rank individuals by relative risk even if their absolute risk estimates were inaccurate. It is therefore important to examine both the model C-statistics and the CHARGE-AF calibration to gain a more holistic understanding of AF predictive ability. (15)

Results also showed a significant interaction between sex and CHARGE-AF score, with a stronger association among females than among males. The interaction is supported by existing literature, which indicates that the same risk factors for AF may affect males and females differently. (18) Prior research also suggests that certain cardiovascular risk factors (such as biomarkers) are more strongly associated with incident AF among women. (19) This interaction warrants further investigation into potential mechanisms underlying the sex-specific differences.

### Strengths and limitations

Our study used a well-established prospective cohort with detailed clinic visit data. (8) We obtained a large sample that was followed for over two decades. Multiple high-quality methods were used to ensure that AF was detected with high sensitivity and specificity. (13) Finally, we evaluated both discrimination and calibration for our models to provide a holistic performance assessment. (14,15)

We also note some limitations. The final analytic sample included only participants who attended each of the first four ARIC clinic visits; this may introduce selection bias (all participants in our analysis are adherent to clinical visits), and results may not be generalizable to all populations. In addition, although many established AF risk factors (i.e., age, smoking, blood pressure, and diabetes) are accounted for in the CHARGE-AF calculation, our Cox regression models did not adjust for covariates beyond those in the risk score itself. Lastly, the CHARGE-AF score does not incorporate circulating biomarkers or genomic data, which may provide additional benefit for risk prediction. (20–22)

## Conclusion

The CHARGE-AF score demonstrated a strong association with incident AF, with a higher score corresponding to increased AF risk. The most recent CHARGE-AF score provided the most predictive value, and incorporating prior clinical information, as summarized by prior values of the risk score, did not meaningfully improve model performance. We also observed significant interaction by sex, with a stronger association between CHARGE-AF and incident AF among females compared to males. Overall, our findings suggest that a single, most recent CHARGE-AF score may be sufficient to estimate AF risk, even when longitudinal risk measurements are available.

## Data Availability

All data produced are available upon reasonable request to the ARIC Coordinating Center.

## Acknowledgments

The authors thank the staff and participants of the ARIC study for their important contributions.

## Funding

The Atherosclerosis Risk in Communities study has been funded in whole or in part with Federal funds from the National Heart, Lung, and Blood Institute, National Institutes of Health, Department of Health, and Human Services, under Contract nos. (75N92022D00001, 75N92022D00002, 75N92022D00003, 75N92022D00004, 75N92022D00005). L.Y.L. is supported by the National Center for Advancing Translational Sciences of the National Institutes of Health under Award Numbers TL1TR002382 and UL1TR002378. The content is solely the responsibility of the authors and does not necessarily represent the official views of the National Institutes of Health.

## Disclosures

None

## Notes

### Competing Interest Statement

The authors have declared no competing interest.

### Author Declarations

IRB of all ARIC participating institutions (University of Minnesota, Johns Hopkins University, University of North Carolina Chapel Hill, Wake Forest University, University of Mississippi Medical Center) gave ethical approval for this work.

## References

1. Lakshminarayan K, Anderson DC, Herzog CA, Qureshi AI. Clinical Epidemiology of Atrial Fibrillation and Related Cerebrovascular Events in the United States. Neurologist. 2008 May;14(3):143–50. doi:10.1097/NRL.0b013e31815cffae PubMed PMID: 18469671; PubMed Central PMCID: PMC5619693.

2. Odutayo A, Wong CX, Hsiao AJ, Hopewell S, Altman DG, Emdin CA. Atrial fibrillation and risks of cardiovascular disease, renal disease, and death: systematic review and meta-analysis. BMJ. 2016 Sep 6;354:i4482. doi:10.1136/bmj.i4482 PubMed PMID: 27599725.

3. Feinberg WM, Blackshear JL, Laupacis A, Kronmal R, Hart RG. Prevalence, Age Distribution, and Gender of Patients With Atrial Fibrillation: Analysis and Implications. Arch Intern Med. 1995 Mar 13;155(5):469–73. doi:10.1001/archinte.1995.00430050045005

4. Alonso A, Krijthe BP, Aspelund T, Stepas KA, Pencina MJ, Moser CB, et al. Simple risk model predicts incidence of atrial fibrillation in a racially and geographically diverse population: the CHARGE-AF consortium. J Am Heart Assoc. 2013 Mar 18;2(2):e000102. doi:10.1161/JAHA.112.000102 PubMed PMID: 23537808; PubMed Central PMCID: PMC3647274.

5. Pfister R, Brägelmann J, Michels G, Wareham NJ, Luben R, Khaw KT. Performance of the CHARGE-AF risk model for incident atrial fibrillation in the EPIC Norfolk cohort. Eur J Prev Cardiol. 2015 Jul;22(7):932–9. doi:10.1177/2047487314544045 PubMed PMID: 25059930.

6. Alonso A, Roetker NS, Soliman EZ, Chen LY, Greenland P, Heckbert SR. Prediction of Atrial Fibrillation in a Racially Diverse Cohort: The Multi-Ethnic Study of Atherosclerosis (MESA). J Am Heart Assoc. 2016 Feb 23;5(2):e003077. doi:10.1161/JAHA.115.003077 PubMed PMID: 26908413; PubMed Central PMCID: PMC4802458.

7. Himmelreich JCL, Lucassen WAM, Harskamp RE, Aussems C, van Weert HCPM, Nielen MMJ. CHARGE-AF in a national routine primary care electronic health records database in the Netherlands: validation for 5-year risk of atrial fibrillation and implications for patient selection in atrial fibrillation screening. Open Heart. 2021 Jan 18;8(1):e001459. doi:10.1136/openhrt-2020-001459 PubMed PMID: 33462107; PubMed Central PMCID: PMC7816907.

8. Project Overview | ARIC [Internet]. [cited 2026 Jan 31]. Available from: https://www5.cscc.unc.edu/aric9/about/project_overview

9. Wright JD, Folsom AR, Coresh J, Sharrett AR, Couper D, Wagenknecht LE, et al. The ARIC (Atherosclerosis Risk In Communities) Study: JACC Focus Seminar 3/8. J Am Coll Cardiol. 2021 Jun 15;77(23):2939–59. doi:10.1016/j.jacc.2021.04.035 PubMed PMID: 34112321; PubMed Central PMCID: PMC8667593.

10. Li LY, Li L, Chen LY, Soliman EZ, Alonso A. The association between alcohol intake and incident atrial fibrillation in older adults: The ARIC cohort. PLoS One. 2024 Nov 21;19(11):e0314207. doi:10.1371/journal.pone.0314207 PubMed PMID: 39570908; PubMed Central PMCID: PMC11581337.

11. Chamberlain AM, Agarwal SK, Folsom AR, Soliman EZ, Chambless LE, Crow R, et al. A Clinical Risk Score for Atrial Fibrillation in a Biracial Prospective Cohort (From the Atherosclerosis Risk in Communities (ARIC) Study). Am J Cardiol. 2011 Jan;107(1):85–91. doi:10.1016/j.amjcard.2010.08.049 PubMed PMID: 21146692; PubMed Central PMCID: PMC3031130.

12. Alonso A, Agarwal SK, Soliman EZ, Ambrose M, Chamberlain AM, Prineas RJ, et al. Incidence of atrial fibrillation in whites and African-Americans: the Atherosclerosis Risk in Communities (ARIC) study. Am Heart J. 2009 Jul;158(1):111–7. doi:10.1016/j.ahj.2009.05.010 PubMed PMID: 19540400; PubMed Central PMCID: PMC2720573.

13. Ghelani KP, Chen LY, Norby FL, Soliman EZ, Koton S, Alonso A. Thirty_-_ Year Trends in the Incidence of Atrial Fibrillation: The ARIC Study. Journal of the American Heart Association. 2022 Apr 19;11(8):e023583. doi:10.1161/JAHA.121.023583

14. Pencina MJ, D’Agostino RB Sr. Evaluating Discrimination of Risk Prediction Models: The C Statistic. JAMA. 2015 Sep 8;314(10):1063–4. doi:10.1001/jama.2015.11082

15. Steyerberg EW, Vickers AJ, Cook NR, Gerds T, Gonen M, Obuchowski N, et al. Assessing the performance of prediction models: a framework for traditional and novel measures. Epidemiology. 2010 Jan;21(1):128–38. doi:10.1097/EDE.0b013e3181c30fb2 PubMed PMID: 20010215; PubMed Central PMCID: PMC3575184.

16. O’Neal WT, Alonso A. The appropriate use of risk scores in the prediction of atrial fibrillation. J Thorac Dis. 2016 Oct;8(10):E1391–4. doi:10.21037/jtd.2016.10.96 PubMed PMID: 27867638; PubMed Central PMCID: PMC5107446.

17. Christophersen IE, Yin X, Larson MG, Lubitz SA, Magnani JW, McManus DD, et al. A comparison of the CHARGE-AF and the CHA2DS2-VASc risk scores for prediction of atrial fibrillation in the Framingham Heart Study. Am Heart J. 2016 Aug;178:45–54. doi:10.1016/j.ahj.2016.05.004 PubMed PMID: 27502851; PubMed Central PMCID: PMC5344697.

18. Zhang F, Hu Y, Wang J, Zhang Q, Yang X, Du Z, et al. Sex Differences in the Association Between Polygenic Risk Score and Atrial Fibrillation Incidence: A Prospective Cohort Study. Can J Cardiol. 2025 Dec;41(12):2554–63. doi:10.1016/j.cjca.2025.09.034 PubMed PMID: 41015248.

19. Peters SAE, Woodward M. Established and novel risk factors for atrial fibrillation in women compared with men. Heart. 2019 Feb;105(3):226–34. doi:10.1136/heartjnl-2018-313630 PubMed PMID: 30158135.

20. Sinner MF, Stepas KA, Moser CB, Krijthe BP, Aspelund T, Sotoodehnia N, et al. B-type natriuretic peptide and C-reactive protein in the prediction of atrial fibrillation risk: the CHARGE-AF Consortium of community-based cohort studies. Europace. 2014 Oct;16(10):1426–33. doi:10.1093/europace/euu175 PubMed PMID: 25037055; PubMed Central PMCID: PMC4197895.

21. Yao Y, Zhang MJ, Wang W, Zhuang Z, He R, Ji Y, et al. Multimodal data integration to predict atrial fibrillation. Eur Heart J Digit Health. 2025 Jan;6(1):126–36. doi:10.1093/ehjdh/ztae081 PubMed PMID: 39846068; PubMed Central PMCID: PMC11750194.

22. Roselli C, Surakka I, Olesen MS, Sveinbjornsson G, Marston NA, Choi SH, et al. Meta-analysis of genome-wide associations and polygenic risk prediction for atrial fibrillation in more than 180,000 cases. Nat Genet. 2025 Mar;57(3):539–47. doi:10.1038/s41588-024-02072-3 PubMed PMID: 40050429; PubMed Central PMCID: PMC12094172.

